# The relationship between ethnicity and Multiple Sclerosis characteristics in the United Kingdom: a UK MS Register study

**DOI:** 10.1101/2024.02.02.24301981

**Authors:** Benjamin M Jacobs, Luisa Schalk, Emily Tregaskis-Daniels, Pooja Tank, Sadid Hoque, Michelle Peter, Katherine Tuite-Dalton, James Witts, UK MS Register Study Group, Riley Bove, Ruth Dobson

**Author notes:** **Correspondence to:** Professor Ruth Dobson, Centre for Preventive Neurology, Wolfson Institute of Population Health, Queen Mary University of London, UK.

## Abstract

Previous studies have suggested differences in Multiple Sclerosis severity between individuals from different ethnic backgrounds. However, these measures of severity have been focussed on markers of physical disability derived from clinical records. We sought to determine the association between self-reported ethnicity and Multiple Sclerosis severity across a range of domains in a prospective cohort based in the United Kingdom. Data were obtained from the United Kingdom Multiple Sclerosis Register, a longitudinal cohort study of >20,000 persons with Multiple Sclerosis. We examined the association between self-reported ethnic background and age at onset, site of onset, and a variety of participant-reported severity measures: the Expanded Disability Status Scale, the Multiple Sclerosis Impact Scale, the Multiple Sclerosis Walking Scale, the Fatigue Severity Scale, and the EuroQol-5D quality of life index. We used multivariable linear regression models adjusted for age, sex, and Multiple Sclerosis subtype (Primary Progressive vs other). We explored the association between ethnicity and impact of Multiple Sclerosis using Cox proportional hazards models to assess the rate of disability progression. Data were available for 14,264 people with Multiple Sclerosis, including 380 participants from self-reported Black (n=151) or South Asian (n=229) ethnic backgrounds. Age at Multiple Sclerosis onset and diagnosis was lower in the participants of South Asian (median onset 31.0) and Black (median onset 33.0) ethnicity compared with White ethnicity (median onset 35.0, n=13,884). We found no statistically-significant evidence for an association between ethnic background on baseline Multiple Sclerosis severity scores in any of the scores tested across a range of sensitivity analyses. In longitudinal analysis, we did not find evidence for an impact of ethnicity on five-year risk of disability progression. In this large, ethnically-diverse Multiple Sclerosis cohort with universal access to healthcare, we find no association between ethnic background *per se* and MS severity, whether in cross-sectional or longitudinal analyses. These findings suggest that other factors, such as socioeconomic status and structural inequalities, may explain previous findings of heterogeneity between ethnic groups.

## Introduction

Multiple Sclerosis (MS) affects individuals from all ethnic backgrounds, running counter to the historical belief that it was predominantly a disease of White individuals^1–9^. It has been suggested that ethnic background may be associated with differences in MS disease course, including age at onset, speed of progression, and response to treatment. Specifically, a number of previous studies have suggested that MS has a more aggressive course in people from Black and Hispanic ethnic backgrounds, with earlier onset, more rapid disability progression, greater cognitive and physical impairment, and poorer response to disease-modifying therapies ^10–21^. Other studies have shown an earlier age at onset in Southeast Asian^22^ and South Asian persons^23,24^.

However, most available data has come from hospital-based cohorts in the United States. Therefore, it remains unclear whether MS severity and progression are associated with any biological aspects of ethnic background *per se*, or whether the observed differences are driven by social determinants of health such as systemic or medical racism, healthcare inequalities, and poorer access to timely diagnosis and treatment among minority ethnic groups^25^, which may differ between countries and healthcare systems. Clarifying whether ethnicity is itself associated with an unfavourable MS prognosis is crucial to further exploration of potential genetic and/or gene-environment interaction pathways responsible for disease progression, or conversely, highlighting the need to address social determinants of health which could predispose to a more aggressive MS disease course.

Answering this question robustly requires prospective data from population-based cohorts, which encompass diversity in terms of ethnicity, geographic location and healthcare provider. There have been no studies examining the relationship between ethnicity and MS severity in large, longitudinal cohorts of this nature to date; and specifically none in countries where universal access to healthcare is routine. The UK MS Register (UKMSR) is a longitudinal cohort study of adults with MS living in the UK, which has recruited over 20,000 people with MS since 2011^26,27^. This resource offers a unique opportunity to study potential contributors to disease severity in a longitudinal cohort. The UKMSR collects a range of participant-reported outcome measures which provide insight into multiple dimensions of disability in MS, including fatigue, mood, and quality of life in addition to traditional scores based largely on physical disability, i.e. the online version of the Expanded Disability Status Scale (EDSS).

In this paper, we use the UKMSR to explore the association between self-reported ethnic background and MS severity measures across a range of domains. We use both patient-reported and disease-specific disability measures in order to probe the impact of MS severity across different ethnic groups, examining both point estimates and longitudinal trajectories of severity and impact.

## Methods

### Cohort

Data were extracted from the UK MS Register (UKMSR) in November 2023. Details of participant recruitment and study design have been previously described in detail^26,27^. In brief, baseline demographic characteristics (including age at onset, recruitment, and self-stated gender), self-reported disease type, treatment details and lifestyle information are gathered at recruitment, and participants are surveyed on a six-monthly basis using a battery of participant-reported outcome measures.

### Variable definitions

#### Demographic variables and ethnicity

Self-reported ethnic background is selected by participants at registration using the UK 2001 Census categories (https://www.nomisweb.co.uk/datasets/c2021ts021) which are reflected in the NHS data dictionary (https://www.datadictionary.nhs.uk/data_elements/ethnic_category.html). Granular categories were condensed into four parent groups (White, Asian, Black, Mixed/other). Other demographic variables obtained via self-report include age at MS symptom onset, age at diagnosis, MS type at onset, disease-modifying therapy (DMT) exposure, educational attainment and gender. DMT exposure was defined using self-reported DMT records indicating the drug had started prior to study enrolment. Participants with no DMT records prior to enrolment were considered unexposed to DMT for the purposes of this study. High-efficacy DMTs were defined as natalizumab, anti-CD20 therapy (ocrelizumab, ofatumumab), S1P inhibitors (fingolimod, sipinomod), cladribine, and alemtuzumab.

#### MS outcome measures

We used five separate outcome measures of MS severity: the online (web) Expanded Disability Scale Score (EDSS); and four participant-reported outcome measures (PROMs): the physical dimension of the Multiple Sclerosis Impact Scale (MSIS29 version 2), the EuroQol 5D-3L/5L Visual Analogue Scale (VAS), the Fatigue Severity Scale, and the MS Walking Scale.

EDSS values were evaluated via an online self-administered EDSS scale (the webEDSS^28^). From this we calculated the age-adjusted EDSS score, the ARMSS (global Age-Related Multiple Sclerosis Severity Score; gARMSS) using the ‘ms.sev’ R package^29,30^. We used the 20 ‘physical’ questions of the MSIS29 version 2 to calculate a normalised summary score as previously described^31^. Briefly, each of the questions in the MSIS29-v2 is scored from 1 to 4, with higher scores indicating greater disability. The sum of the 20 questions in the MSIS29 physical domain therefore ranges from 20 to 80. We normalised these scores as follows:

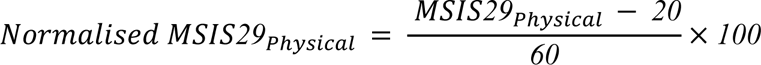

This procedure yielded normalised MSIS29_Physical_ scores ranging from 0 - 100, with higher scores indicating worse MS. The EQ5D VAS is a self-rating of current health-related quality of life from 0 - 100, with higher scores indicating better quality of life. The fatigue severity score (FSS) consists of seven questions rated from 1 to 9, with 9 indicating a higher degree of impairment due to fatigue. The total score ranges from 9 to 63, with higher scores indicating worse fatigue – these scores were normalised to range from 0 - 100 using the same procedure as for the MSIS^31^,:

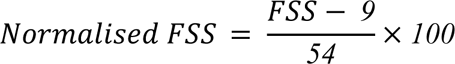

The MS Walking Scale (MSWS) consists of twelve questions evaluating gait which are rated from 1 to 5, with higher scores indicating more severe disability. The raw scores therefore sum to between 12 and 60. Again, these score were normalised to range from 0 - 100 as follows^32^:

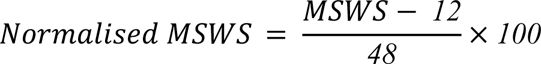

From this point we refer to the normalised physical MSIS score, the normalised FSS, and the normalised MSWS as the MSIS, FSS, and MSWS respectively.

### Participant inclusion/exclusion criteria

#### Primary analysis (cross-sectional) cohort

We considered an initial dataset of 19,153 participants enrolled in the UKMSR with complete demographic data (age, sex, MS type at onset, age at MS symptom onset and diagnosis, ethnicity, and year of birth; table 1; figure 1). We then excluded those diagnosed prior to the 2001 McDonald criteria (remaining n=14,580). We restricted the dataset to the 14,264 participants who identified as one of the following ethnic groups: “I am Asian or British Asian (Indian / Pakistani / Bangladeshi”, “I am Black or Black British (Caribbean, African, Other)”, “I am white (British, Irish, Other)”, referred to from this point onwards as ‘South Asian’, ‘Black’, ‘White’ respectively.

**Figure 1:**
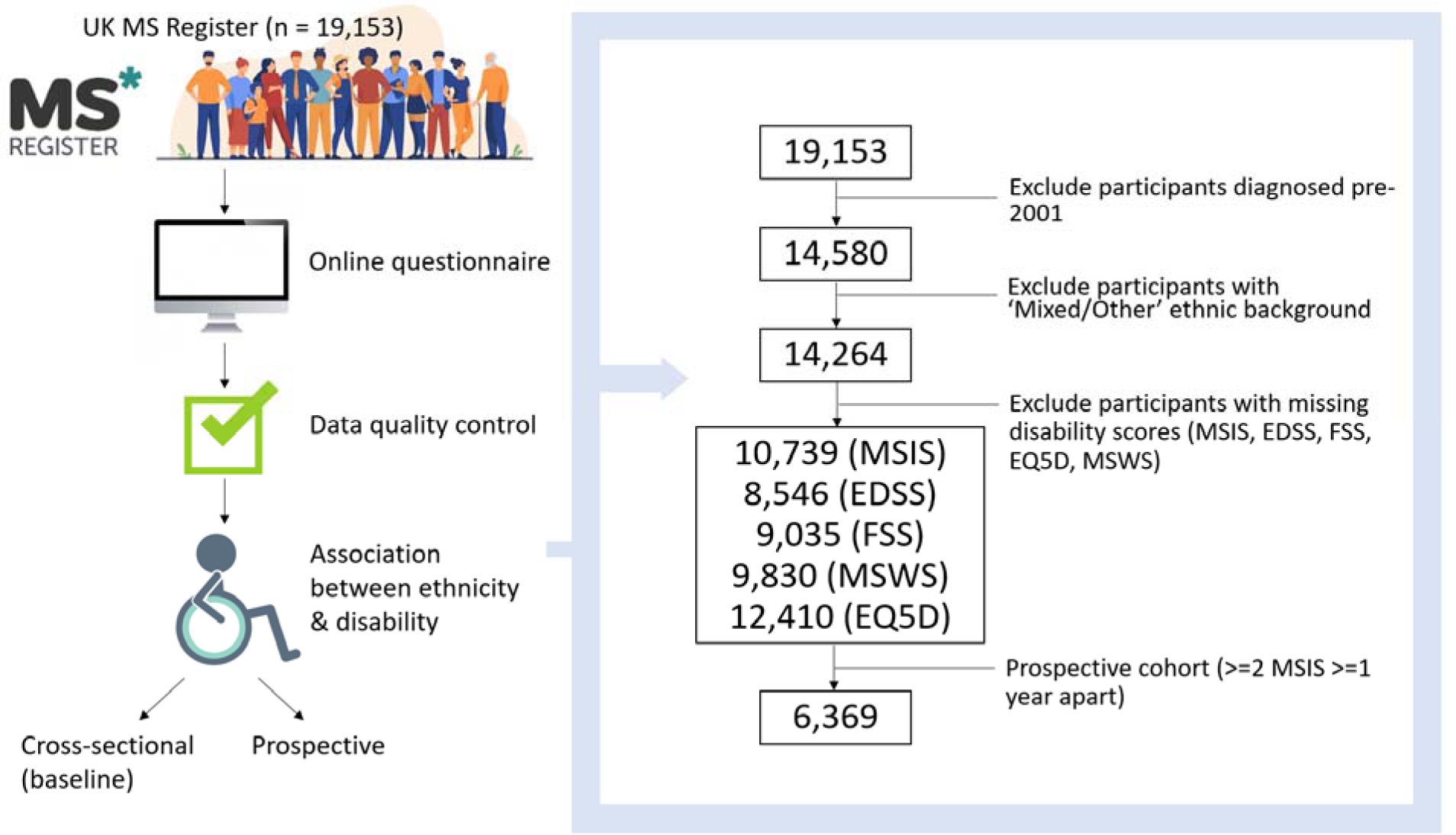
flowchart depicting the experiment design, including participant inclusion/exclusion criteria. Participants from Black (n=151) and South Asian (n=229) ethnic backgrounds reported younger age at both symptom onset and diagnosis than White participants (figure 2, table 1, Kruskal-Wallis *P* < 0.001). The median age of symptom onset was 31.0 in South Asian participants, 33.0 in Black participants, and 35.0 in White participants. The difference in age at diagnosis was even more striking, with South Asian participants diagnosed on average seven years earlier than White participants (median 33.0 vs 40.0).

**Figure 2:**
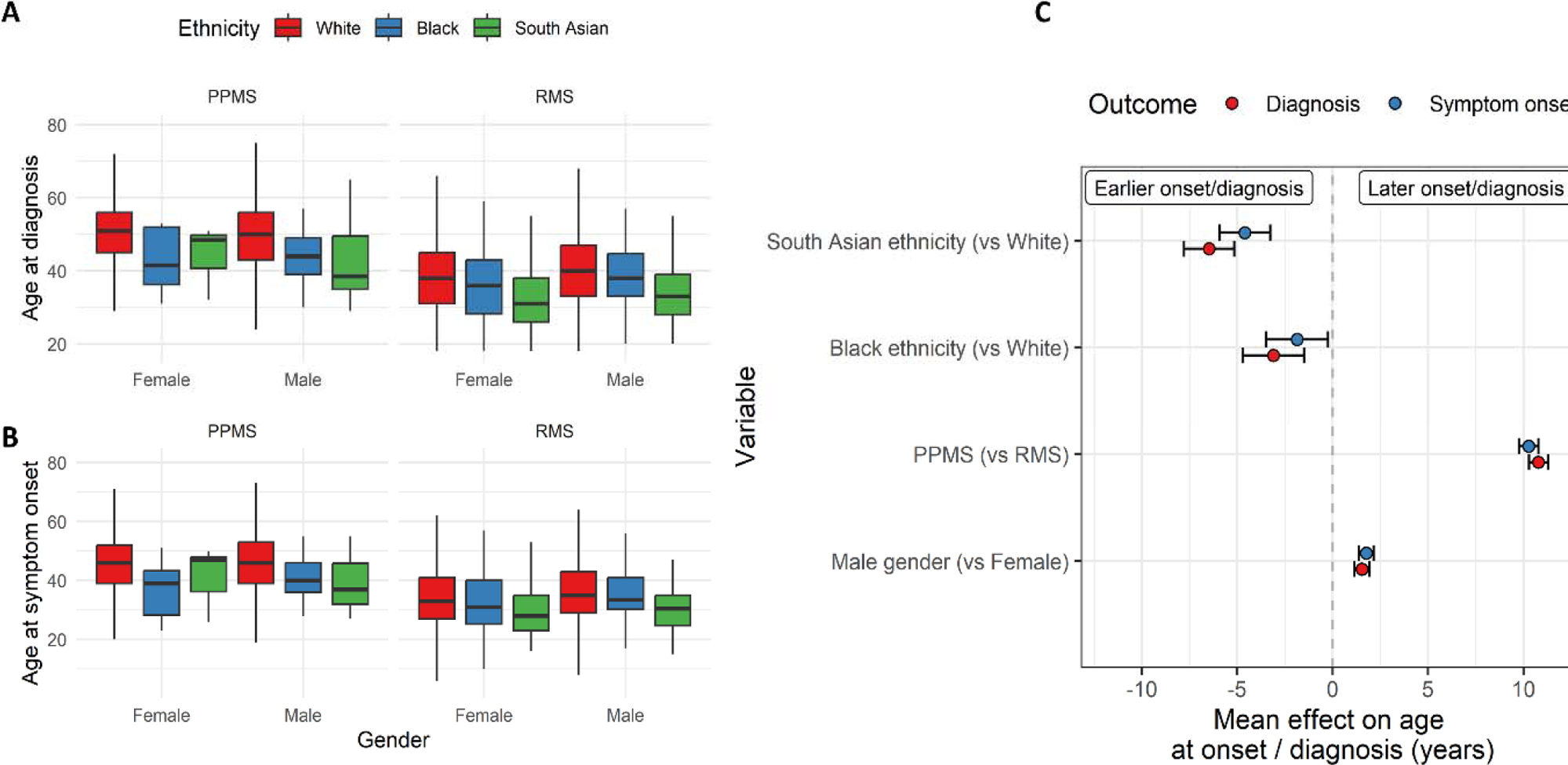
age at MS onset and diagnosis is younger in South Asian and Black participants. A – boxplots showing the age at diagnosis (A) and symptom onset (B) split by ethnicity, MS subtype, and gender. C – regression coefficients from linear models estimating the impact of ethnicity, MS subtype, and gender on MS onset and age at diagnosis. Points represent the beta coefficient +/− 95% CI, i.e. the estimated mean difference in age at diagnosis/onset given each variable. The dotted line indicates the null. Red points represent the impact on age at diagnosis, and blue points indicate the impact on age at symptom onset. PPMS - primary progressive MS. RMS - Relapse-onset MS.

**Table 1:** characteristics of the cohort.

|  | White | Black | South Asian | P |
| --- | --- | --- | --- | --- |
|  | N=13884 | N=151 | N=229 |  |
| Age at diagnosis | 40.0 [32.0;48.0] | 38.0 [30.0;45.0] | 33.0 [27.0;40.0] | <0.001 |
| Year of diagnosis | 2012 [2007;2016] | 2013 [2009;2017] | 2014 [2010;2017] | <0.001 |
| Gender: |  |  |  | <0.001 |
| Female | 10446 (75.2%) | 111 (73.5%) | 145 (63.3%) |  |
| Male | 3438 (24.8%) | 40 (26.5%) | 84 (36.7%) |  |
| University education: |  |  |  | <0.001 |
| No | 7019 (56.5%) | 69 (51.9%) | 54 (26.2%) |  |
| Yes | 5403 (43.5%) | 64 (48.1%) | 152 (73.8%) |  |
| Age at symptom onset | 35.0 [28.0;44.0] | 33.0 [27.0;41.0] | 31.0 [25.0;37.0] | <0.001 |
| DMT prior to baseline: |  |  |  | 0.021 |
| No | 10826 (78.0%) | 131 (86.8%) | 185 (80.8%) |  |
| Yes | 3058 (22.0%) | 20 (13.2%) | 44 (19.2%) |  |
| PPMS: |  |  |  | 0.901 |
| No | 11269 (86.3%) | 124 (85.5%) | 190 (87.2%) |  |
| Yes | 1786 (13.7%) | 21 (14.5%) | 28 (12.8%) |  |
| Baseline EDSS | 4.50 [3.00;6.50] | 4.50 [3.00;6.50] | 3.75 [2.00;6.50] | 0.050 |
| Age at baseline EDSS | 50.2 [41.4;57.7] | 47.8 [36.3;54.6] | 42.8 [35.2;49.9] | <0.001 |
| Baseline gARMSS | 6.25 [4.10;7.89] | 6.53 [4.92;8.47] | 6.48 [4.27;8.42] | 0.063 |
| Baseline MSIS | 38.3 [16.7;61.7] | 37.5 [15.0;61.7] | 35.0 [15.8;62.5] | 0.978 |
| Age at baseline MSIS | 47.6 [39.2;55.4] | 45.5 [36.3;52.0] | 39.8 [33.8;47.0] | <0.001 |

In the cross-sectional analysis of MS severity measures, we obtained the earliest participant-reported severity outcome measure in each domain (webEDSS, MSIS, FSS, MSWS, and EQ5D) for each participant (n for each outcome shown in figure 1).

#### Longitudinal cohort

To assess the relationship between ethnicity and long-term disability progression in a longitudinal setting, we defined a longitudinal cohort by identifying participants from the cross-sectional cohort who met the following criteria:

1. At least two valid MSIS recordings - the earliest (baseline) reading and at least one reading and within 5 years of the baseline.
2. Baseline MSIS <=90.
3. Follow-up reading >=1 year from the baseline

From the 8,315 participants with a baseline MSIS and at least one follow-up MSIS reading, 6,660 had readings at least a year apart. We restricted follow-up to five years due to differential rates of missing data between ethnic groups after this time point and subsequent risk of bias due to informative censoring, leaving 6,469 participants. We excluded 100 participants whose baseline MSIS was greater than 90 (i.e. they could not experience the outcome of a 10-point increase as the score goes up to 100), providing a final analysis cohort of 6,369 participants.

### Statistical analysis

#### Comparison of demographic characteristics

To compare demographic baseline characteristics between groups, we used Kruskal-Wallis tests (for continuous variables) and chi-squared tests (for categorical variables). Unless specified, categorical variables are reported as n (%) and continuous variables as median (interquartile range).

#### Association of ethnicity with clinical characteristics

We tested for association between ethnicity and age at MS onset and diagnosis using linear regression models. Models were adjusted for gender and progressive-onset disease. Visual inspection of the distributions of both age at symptom onset and age at diagnosis confirmed both were normally-distributed. Inspection of models confirmed that linear regression assumptions of linearity and normally-distributed variances were satisfied. After excluding the small number of participants (n=31) reporting an MS diagnosis prior to their reported symptom onset, we calculated the lag from symptom onset to diagnosis, we derived an estimated time from onset to diagnosis as the difference between the stated age at onset and age at diagnosis. As this variable was not normally-distributed, we tested for association with ethnicity using bootstrapped linear regression models adjusted for gender and progressive-onset disease. We explored the association between ethnicity and site of onset using multinomial logistic regression models adjusted for gender and progressive-onset disease.

#### Association of ethnicity with baseline severity outcome measures

Associations between ethnicity and baseline MS severity outcome measures were tested using linear regression models, with the ‘White’ ethnic group the reference category. For the primary analyses, regression models were adjusted for the age at the time of recording, gender, and progressive-onset disease. Age at symptom onset and diagnosis were not included in the models due to collinearity as described below. The outcome measure (dependent variable) for these regression models was the earliest raw (i.e. untransformed) severity score per participant. Participants who did not have any recorded values for a particular severity score were excluded from the relevant regression model.

The distribution of severity measure scores was not normal, thus violating the assumptions of linear regression. We therefore calculated the confidence intervals, standard errors, and P values of regression coefficients with bootstrapping. For each model, we resampled the dataset with replacement and refitted the regression model with the resampled data. This procedure was repeated 1,000 times for each model. 95% confidence intervals were derived from the empirical quantiles of the sampling distribution. As the sampling distribution for the regression coefficients was normally distributed, we calculated two-tailed P values for the null hypothesis that beta = 0 as (beta - mean(beta))/ sd(beta). We confirmed the expected strong correlations between baseline severity measures. Regression models were inspected to ensure that the linearity assumption was met. We inspected the correlation between predictors to look for evidence of multicollinearity (see above). The high correlation between age at symptom onset, age at diagnosis, and age at the time of severity measure recording (supplementary measure) prompted us to use only one of these predictors (age at test) in the primary analysis.

To account for demographic and disease-related differences between ethnic groups in the UKMSR which could confound the interpretation of MS outcome measures, we created a matched cohort in which each participant reporting Black or South Asian ethnicity was matched to exactly two participants reporting White ethnicity. Matching was performed on age at diagnosis (rounded to the nearest year), year of diagnosis (rounded to the nearest year), gender, and MS subtype (relapse-onset vs primary-progressive disease). Regression models were applied to this matched dataset in the same fashion as for the primary analysis.

#### Association of ethnicity with longitudinal severity outcome measures

We examined the relationship between ethnicity and longitudinal disability progression using the MSIS-29 physical subscale. We filtered the cohort to the 6,369 participants with longitudinal MSIS readings (defined above). For the time-to-event analysis we defined ‘disability progression’ as the occurrence of an MSIS score 10 points greater than the baseline score, as this has been demonstrated to represent a clinically-meaningful MSIS increment^33^. To distinguish sustained disability progression from transient worsening, which could be a result of relapse-associated worsening, measurement error, or pseudo-relapse, we identified those with a further follow-up MSIS at least three months later.

The time-to-event was defined as the time from the baseline score to the first MSIS score 10 points above baseline. If participants did not experience disability progression during the follow-up period they were censored, with the censoring time defined as the last (i.e. most recent) MSIS score. We used Cox proportional hazards regression to model theeffect of ethnicity of sustained disability progression. In the primary analysis we adjusted for age at baseline, baseline MSIS, gender, and progressive-onset disease. Cox models were inspected for linearity of numerical predictors, validity of the proportional hazards assumption, and influence of individual observations.

We conducted several sensitivity analyses. First, we adjusted for the impact of disease-modifying treatment. Second, we used a more lenient definition of progression, including those with only one MSIS score 10 points above their baseline (i.e. not stipulating the presence of a follow-up score as we did to define sustained progression). Third, we repeated the analysis in the matched cohort defined in the previous section, i.e. a subset of the cohort in which each Black or South Asian participant was matched to exactly two participants of White ethnicity on age at diagnosis, year of diagnosis, gender, and MS subtype.

### Power calculations

We performed empirical power calculations for the cross-sectional (baseline) analysis by generating simulated datasets of the same size as the ethnic groups in the study (White n = 13,884, Black n=151, South Asian n=229). We simulated a normal distribution of MSIS scores as a simplification, using the same standard deviation (26) as that observed in the whole population. For the White reference group we used the observed population mean of 40. We truncated these distributions such that randomly-generated scores outside of the 0-100 range were resampled, i.e. these were truncated normal distributions. We performed the same procedure for the non-reference group, altering the mean of the distribution, i.e. the true population difference between groups. We estimated power at alpha < 5% as the proportion of 1,000 bootstrap iterations in which the linear regression model Wald test P value term was <0.05.

For the survival analysis, we performed empirical power calculations by simulating survival times (i.e. time to a 10-point step change in MSIS score) using exponential distributions with a fixed hazard of 0.1 (i.e. 1 event per 10 person-years) for the reference (White) group, which closely resembled the observed data. We used the same N values for the reference (White) and non-reference groups as for the cross-sectional analysis. We adjusted the hazard for the non-reference group by multiplying the reference hazard by a range of hazard ratios, from 1 (no effect) to 2 (a doubling of the hazard). Over 1,000 bootstrap iterations, we performed unadjusted Cox regression and calculated empirical power as the proportion of iterations with a P value of < 0.05.

### Computing, data and code availability

All analyses were conducted in R version 4.1.3 within the UKMSR secure research environment (UKSERP)^27^. All code used in these analyses is available at https://benjacobs123456.github.io/ukmsr_ethnicity/. Access to UKMSR data is open to all researchers on application. Details of how to apply for the data can be found here: https://ukmsregister.org/Research/WorkingWithUs.

### Ethical approval

The UK Multiple Sclerosis Register has research ethics approval from South West Central Bristol Research Ethics Committee initially as 16/SW/0194 currently 21/SW/0085. This project was approved by the UKMSR Scientific Steering Committee.

## Results

### Ethnicity correlates with age of Multiple Sclerosis onset

The UKMSR cohort (n=14,264 with complete demographic data) was broadly representative of the UK MS population, with a female predominance (n=10,702, 75.0%), primarily relapse-onset disease (n=11,583, 81.2%), largely identifying as White (n=13,884, 97.3%, comparable to our previous population-based study^23^), with median age at diagnosis of 40.0 (IQR 16), and median age at symptom onset of 35.0 (IQR 15). While the gender ratio appeared similar between White (n=10,466, 75.2% female) and Black (n=111, 73.5% female) ethnic groups, the female preponderance was less pronounced among South Asian participants (n=145, 63.3% female). The proportion of participants with self-reported Primary Progressive MS was similar across ethnic groups (table 1).

The association between Black or South Asian ethnicity and earlier MS onset persisted despite adjustment for gender and progressive-onset disease. These models reproduced the expected finding that male gender and progressive-onset disease were associated with later symptom onset and diagnosis. Restricting the analysis to participants diagnosed after 2010, when revisions to the diagnostic criteria made it simpler to diagnose MS based on a single scan (and would therefore plausibly lower the average age at diagnosis) yielded the same findings^34^.

In contrast to recent data from other cohorts, we did not find evidence for delayed diagnosis among participants from minoritised ethnic groups - the lag from symptom onset to diagnosis was less pronounced in Black (median 2.0 years) and South Asian (median 2.0 years) participants compared with White participants (median 3.0 years). As the distribution of lag times was not normally distributed we adjusted for gender and progressive onset disease using bootstrapped linear regression models. These models confirmed evidence for less diagnostic lag (i.e. shorter time from symptom onset to diagnosis) in the Black (*P* = 0.003) and South Asian (*P* = 0.001) groups.

Around one third of the cohort reported their first MS symptom type (n = 4216, 29.6%), including 39 Black, 68 South Asian and 4109 White participants. Overall sensory symptoms were the most common presenting feature, reported in 1312 participants (31.1% of those with a recorded symptom). Multinomial regression models did not reveal any specific symptom or constellation of symptoms which were associated with ethnic background, although these estimates were imprecise owing to the small numbers (**figure 3**).

**Figure 3:**
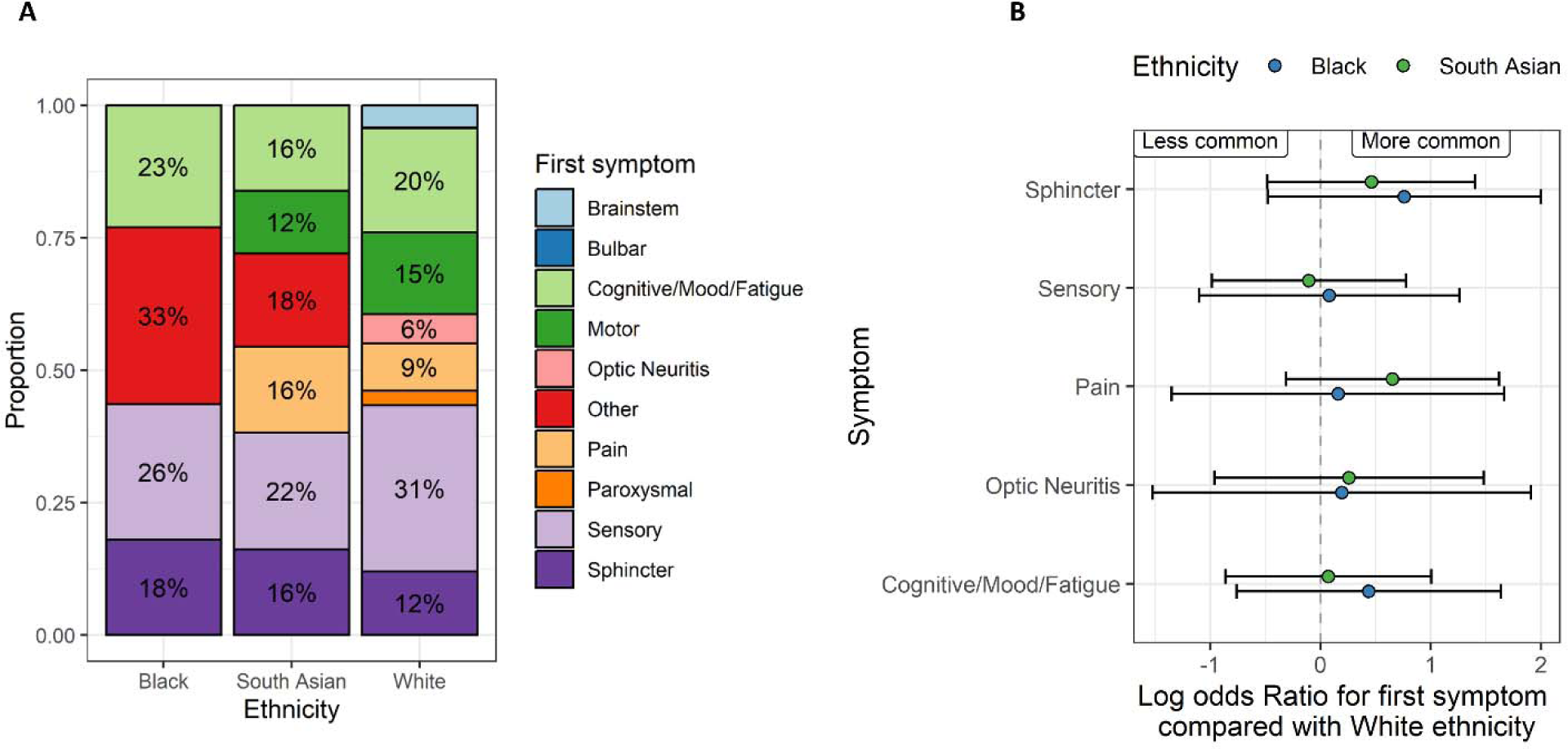
symptom at MS onset does not differ by ethnicity. A – stacked barplots showing the proportion of people in each ethnic group with the presenting symptom denoted by the colour. Note that the denominator is the number of participants with any reported symptom, which was around 1/3 of the cohort. Counts <5 are suppressed and subsumed into the ‘other’ category in the plot. B – regression coefficients showing the results of multinomial logistic regression models. In these models, the point estimates refer to the log odds ratio for each symptom compared with motor symptoms at onset, contrasting with the White ethnic group. Beta coefficients and 95% Cis are shown. Symptoms to the right of the null line can be interpreted as ‘more likely to occur as the presenting symptom’ than in the White group, and vice-versa for those to the left of the null. None of these associations achieved statistical significance at a P value of < 0.05. Points are coloured by the ethnic group.

### MS severity measures do not differ by ethnicity

The baseline age-adjusted EDSS scores (gARMSS) were similar across ethnic groups (medians 6.25, 6.5, and 6.5 in the White, South Asian, and Black groups respectively) (table 1). In models adjusted for age, gender, and progressive-onset disease, there was no evidence (*P* < 0.05) for an association between Black or South Asian ethnicity and any of the MS severity scores we studied (**figure 4**). Empirical power calculations suggested that we would have >90% power to detect a clinically-meaningful difference of >10 on the normalized MSIS scale between groups for both the Black and South Asian groups compared with those of White ethnicity.

**Figure 4:**
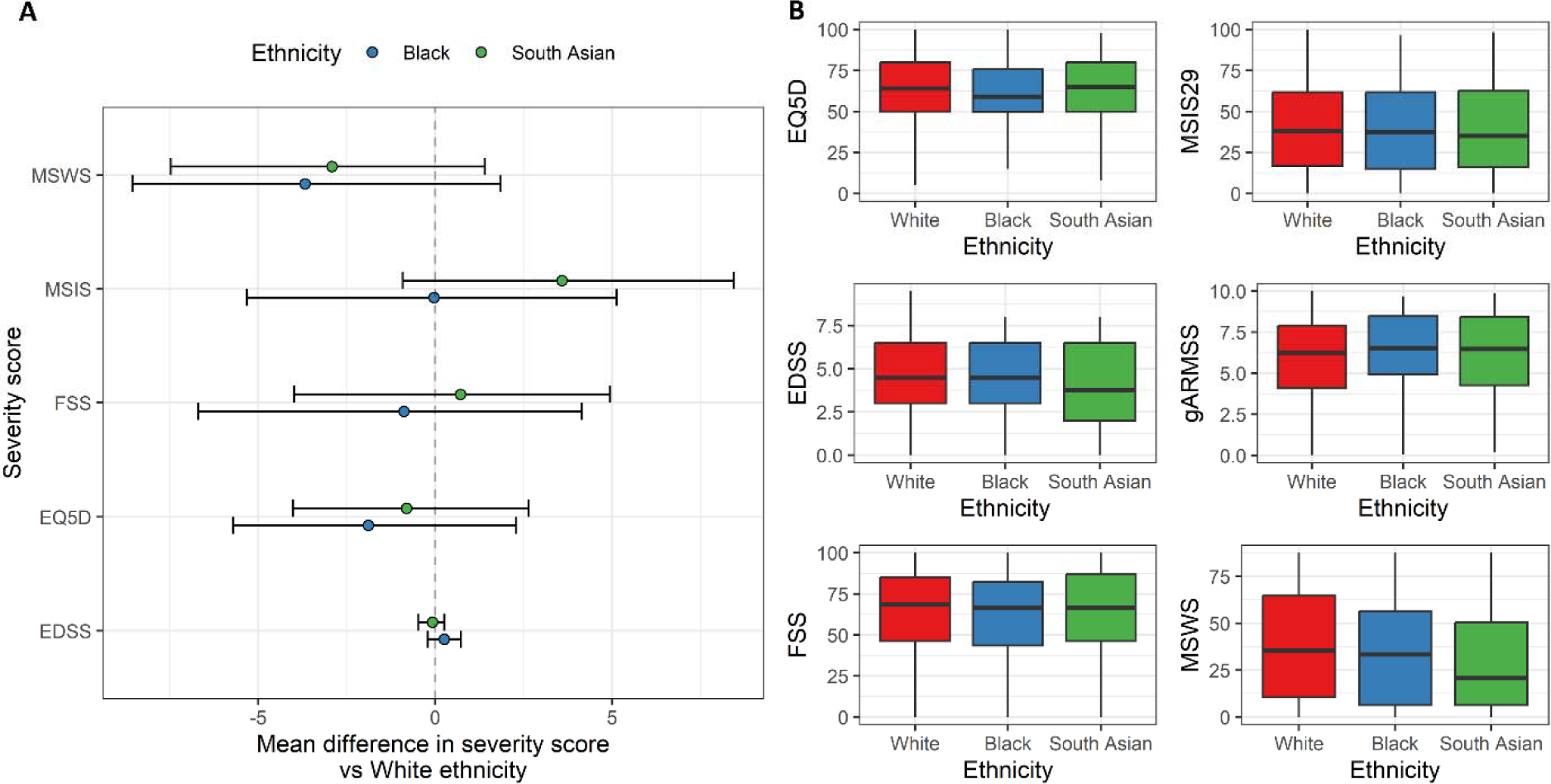
no evidence for association between ethnicity and baseline MS severity measures. A – forest plot showing the regression coefficients and 95% CIs from bootstrapped linear regression models examining the impact of ethnic group on baseline severity scores. Note that all scores except for EQ5D are orientated such that higher value equates to ‘worse’ disease, whereas for EQ5D higher scores indicate higher quality of life. Also note that these values are on the original scale, i.e. the EDSS scale runs from 0-10 whereas the other scores run from 0-100. B – boxplots showing the raw distributions of these severity scores in each ethnic group. In addition, the global ARMSS score – the age-related EDSS – is also shown.

We confirmed the expected associations between progressive-onset disease, male sex, increasing age and worse measures of MS severity. We also observed a consistent protective effect of both prior DMT exposure and university education, consistent with the known benefits of DMT, and secondly the influence of social determinants of health on MS outcomes, as university education is a surrogate for higher socio-economic status.

We did not find any statistically-significant evidence for association between ethnicity and MS severity measures in sensitivity analyses adjusting for age at diagnosis, year of diagnosis, exposure to high efficacy DMT, and educational attainment. Sensitivity analysis using the matched cohort in an attempt to further mitigate bias caused by unbalanced availability of confounders between ethnic groups reinforced the previous findings, with no statistically-significant relationships between ethnicity and any MS severity score.

### Five-year disability progression is not associated with ethnic background

We identified 3,078 participants who had at least one follow-up MSIS greater than 10 points above their baseline score during the five years of follow-up. Of these, 1,966 (63.9%) had sustained disability progression (i.e. another MSIS greater than 10 points above baseline), 594 (19.3%) had non-sustained progression (i.e. other MSIS scores, but none greater than 10 points above baseline), and 518 (16.8%) did not have a further follow-up score. Participants without a further follow-up score were classified as non-progressors for the primary analysis. The median time to disability progression or censoring of 2.4 years, with a maximum follow up time of five years.

Neither Black nor South Asian ethnicity was associated with sustained disability progression over five years of follow-up when compared to those of White ethnicity (HR_Black_ 1.2, 95% CI 0.8 - 2.0; HR_South Asian_ 0.8, 95% CI 0.5 - 1.4) (figure 5a). Factors associated with disability progression were male gender, progressive-onset disease, older age at baseline, and a lower baseline MSIS score (figure 5b). We did not find any evidence for a relationship between ethnicity and disability progression across a range of sensitivity analyses, including in an unadjusted model, with adjustment for high-efficacy DMT exposure (which showed the expected protective effect of high-efficacy DMT - HR 0.7, 95% CI 0.5 - 0.9*, P* = 0.001), with adjustment for age at diagnosis, or in the matched cohort (figure 5). Empirical power calculations indicated that, given these sample sizes, we would expect >80% power to detect a 40% increase in the hazard (HR =1.4) in either South Asian or Black ethnic groups.

**Figure 5:**
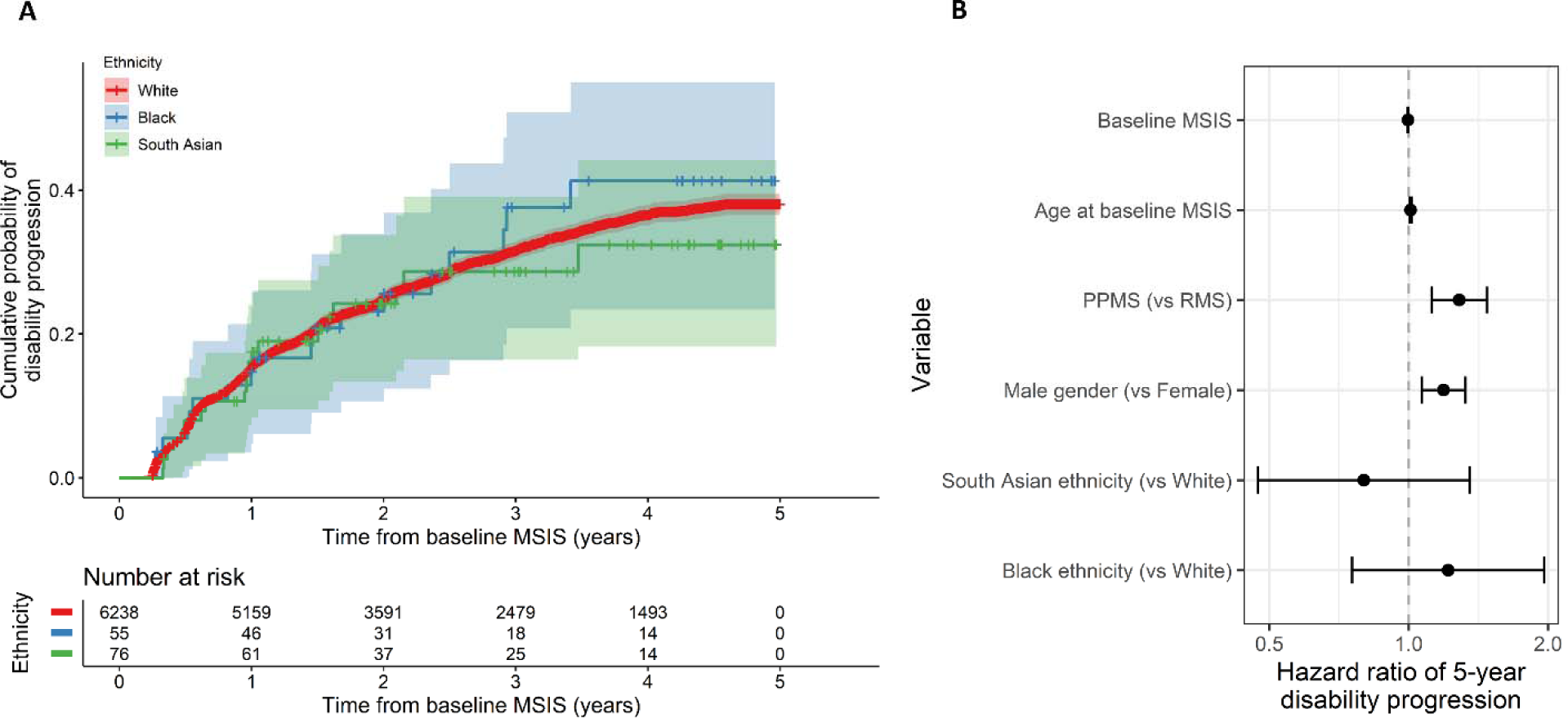
no evidence for association between ethnicity and 5-year disability progression. A – survival curves showing the impact of ethnic group on 5-year disability progression as measured by a 10-point increase in the MSIS29 physical subscale. The x axis indicates time in years from the baseline MSIS measurement, i.e. the earliest recorded measurement for each individual. The curves show the cumulative probability of disability progression. Censored individuals are indicated with crosses. 95% confidence intervals are shown. B – Hazard ratios (and 95% CIs) for disability progression given each factor listed on the y axis. These HRs were derived from a Cox proportional hazards model. PPMS and male gender were associated with higher risk of disability progression, but we did not find any association between ethnic group and progression.

## Discussion

Clarifying whether and how ethnicity impacts on the clinical phenotype of Multiple Sclerosis is important for understanding the determinants of disability, providing accurate prognostic information, and identifying possible healthcare disparities, such as unequal access to timely diagnosis, support, and highly effective disease modifying therapies. We used data from a large longitudinal cohort study - the UK MS Register - including 380 individuals from Black and South Asian ethnic backgrounds, and showed that these individuals tend to be diagnosed with MS at an earlier age, but do not appear to have a more severe disease course. We did not find evidence of a relationship between ethnicity and severity across a range of participant-reported outcomes ascertained at baseline, nor in a longitudinal analysis of sustained disability progression.

Our finding that age at MS onset is earlier in people of Black and South Asian ethnic groups is consistent with our recent UK population-based study of 9,662 MS cases (2.9% South Asian, 2.6% Black) from the Clinical Practice Research Datalink^23^, with baseline phenotype data from our genetic cohort study of ancestrally-diverse people with MS^35^, and with other UK studies^8,24,36^. A primary care records study in East London found that age of onset was earlier in people of South Asian ethnicity but not Black ethnicity people compared with White patients^8^. Data from the United States have been inconsistent regarding age of onset in Black individuals, with some studies showing earlier onset and some later^5,37^. Taken together, these findings suggest that people of South Asian ethnicity living in the UK do experience a younger age of onset, and while this may also be true for people of Black ethnicity living in the UK, the difference may be less stark. The lack of association in our study between ethnicity and cross-sectional disability measures or longitudinal disability progression is reassuring in the context of previous data - largely from the US - showing higher radiological burden, faster progression, and decreased time to fixed disability endpoints among Black individuals with MS^10–21,37^. The available UK data from small cross-sectional studies has suggested higher rates of disability among Black British MS cases, but not among British South Asian cases^24,38^. Our findings are broadly in line with a recent large-scale analysis of >50,000 patients from the MSBase cohort^39^, which found no association between ethnicity and risk of conversion to Secondary Progressive Multiple Sclerosis.

An important caveat in interpreting these results is that ethnicity is not in itself a biological concept. It is often used as a crude proxy for genetic ancestry, but in reality the association between self-reported ethnicity and genetic ancestry is loose. Because of the vagueness inherent in this concept, direct comparison of results between different cohorts is extremely challenging – however studies such as this are useful for understanding the interaction between ethnicity and MS-related outcomes in particular settings. These insights may be specific to the country, healthcare system, and population being studied, but when considered together provide vital evidence into potentially modifiable prognostic factors. Social and healthcare inequalities are a major public health concern in both the UK^40^ and the USA^41^ and may explain a large proportion of the reported differences in MS outcomes between ethnic groups. However, differences in the precise demographics of the UK and the US populations and differences in the healthcare system (insurance-based in the USA; universal access in the UK) mean that direct comparisons between studies are not particularly helpful. Our results should therefore be carefully contextualised – they represent findings from a voluntary, UK-based cohort with universal access to healthcare who are able to self-monitor via the internet, and who are engaged with research.

The key strengths of this work are the richness of the phenotyping, which spans multiple participant-reported outcome measures ascertained over time, and the relatively large sample size. The consistent lack of a relationship between ethnicity and severity across scores for different dimensions and domains of MS-related impairment, both visible (walking) and invisible (fatigue), in both cross-sectional and prospective analysis, and in a range of sensitivity analyses accounting for various possible confounders lends confidence to the null result we report.

The major limitation of this study is the risk of bias due to the voluntary nature of participation in the UK MS Register. Participation in the UKMSR is influenced by a variety of factors including socio-economic status, disability, relationship with healthcare professionals and the healthcare system. These factors may, in theory, skew the cohort towards a relatively less disabled population, and may mean that we are unable to detect a true association between ethnicity and greater disability. In reality, it appears the bias may operate in the other direction - i.e. this population may be more skewed towards a more disabled population with established disease. Regardless of the direction of this bias, relying on voluntary signups rather than population-based recruitment will attract participants who are engaged with healthcare and with research, which will introduce confounding in several domains, including socio-economic status and access to timely diagnosis and treatment.

Although this selection bias does not invalidate the findings, it does mandate replication in a population-based cohort where participation is not influenced by the outcome under study. Compared with a population-based UK MS cohort, the proportion of Black and South Asian cases was lower in the UKMSR (1.1% Black & 1.6% South Asian in UKMSR vs 2.6% Black & 2.9% South Asian in CPRD^23^), suggesting a degree of under-representation of diverse ethnic groups and therefore potential bias. However there is some evidence to suggest that the UKMSR cohort is broadly representative of the MS population in the UK. First, we observe a wide distribution of EDSS scores, including many individuals at the upper end of the disability scale, suggesting that the distribution has not been artificially curtailed. In addition it is reassuring that we are able to replicate known factors associated with greater MS severity, such as progressive-onset disease and older age. We also aim to mitigate confounding by socio-economic status by controlling for educational attainment - although an imperfect proxy, this analysis provides some further support for the results we report.

Other limitations of these data include the exclusive reliance on self-reported metrics, the challenge of properly accounting for the effects of DMT, the possibility of residual confounding by factors such as socio-economic background, and the restriction to only five years of follow-up in the longitudinal analysis. While we attempt to mitigate confounding by performing a wide range of sensitivity analyses adjusting for putative confounders, residual confounding is likely.

Overall these findings suggest that, while individuals from Black and South Asian ethnic backgrounds tend to develop MS and be diagnosed at a younger age, there is no evidence of an association between ethnic background and MS severity in a large UK longitudinal cohort, the UK MS Register. These findings need to be replicated in population-based UK cohorts to mitigate the problem of selection bias. In addition it would be valuable to complement this kind of quantitative approach with qualitative and social science work examining how the road to a diagnosis and the experience of living with MS may differ between people of different ethnic backgrounds. There remains work to be done to understand why certain cohorts show a strong association between ethnicity and disability, an effect we were unable to replicate. This may relate to factors specific to our cohort - in particular its voluntary nature - or alternatively it may reflect the wider social and healthcare environments.

## Data Availability

All analyses were conducted in R version 4.1.3 within the UKMSR secure research environment (UKSERP)27. All code used in these analyses is available at https://benjacobs123456.github.io/ukmsr_ethnicity/. Access to UKMSR data is open to all researchers on application. Details of how to apply for the data can be found here: https://ukmsregister.org/Research/WorkingWithUs.

## Appendix 1

We would like to acknowledge the UK MS Register Research Group collaborators: Alasdair Coles, Jeremy Chataway, Martin Duddy, Hedley Emsley, Helen Ford, Leonora Fisniku, Ian Galea, Timothy Harrower, Jeremy Hobart, Huseyin Huseyin, Christopher M Kipps, Monica Marta, Gavin V McDonnell, Brendan McLean, Owen R Pearson, David Rog, Klaus Schmierer, Basil Sharrack, Agne Straukiene, David V Ford.

